# Leveraging Natural Language Processing and Geospatial Time Series Model to Analyze COVID-19 Vaccination Sentiment Dynamics from Tweets

**DOI:** 10.1101/2022.08.26.22279278

**Authors:** Jiancheng Ye, Jiarui Hai, Zidan Wang, Chumei Wei, Alan Jiacheng Song

## Abstract

**Objective:** To develop and apply a natural language processing (NLP) – based approach to analyze public sentiments on social media and their geographic pattern in the United States toward COVID-19 vaccination. We also provide insights to facilitate the understanding of the public attitudes and concerns regarding COVID-19 vaccination.

**Methods:** We collected Tweet posts by the residents in the United States after the official dissemination of the COVID-19 vaccine. We performed sentiment analysis based on the Bidirectional Encoder Representations from Transformers (BERT) and qualitative content analysis. Time series models were leveraged to describe sentiment trends. Key topics were analyzed longitudinally and geospatially.

**Results:** A total of 3,198,686 Tweets related to COVID-19 vaccination were extracted from January 2021 to February 2022. 2,358,783 Tweets were identified to contain clear opinions, among which 824,755 (35.0%) expressed negative opinions towards vaccination while 1,534,028 (65.0%) demonstrated positive opinions. The accuracy of the BERT model was 79.67%. The key hashtag-based topics include Pfizer, breaking, wearamask, and smartnews. The sentiment towards vaccination across the states showed manifest variability. Key barriers to vaccination include mistrust, hesitancy, safety concern, misinformation, and inequity.

**Conclusion:** We found that opinions toward the COVID-19 vaccination varied across different places and over time. This study demonstrates the potential of an analytical pipeline, which integrates NLP-enabled modeling, time series, and geospatial analyses of social media data. Such analyses could enable real-time assessment, at scale, of public confidence and trust in COVID-19 vaccination, help address the concerns of vaccine skeptics, and provide support for developing tailored policies and communication strategies to maximize uptake.

## INTRODUCTION

In December 2020, the Food and Drug Administration (FDA) in the United States issued the first emergency use authorization (EUA) for use of the COVID-19 vaccine in persons aged 16 years and older for the prevention of COVID-19.[1] As vaccines against COVID-19 are rolled out, there is a pressing need to better understand and monitor public sentiments and address the concerns of vaccine skeptics. This urgency has been exacerbated by the current situation of the global pandemic and growing pressures on health services. Social media, such as Twitter, has been an appropriate source for analyzing public attitudes towards the COVID-19 vaccination.[2]

Artificial intelligence (AI), such as machine learning and natural language processing (NLP), can enable real-time analysis of structured and unstructured data on social media, including public attitudes, demographic determinants, and popular topics.[3] This analysis offers the opportunity to track the dynamic public sentiments and develop proactive communication strategies. In addition, an iterative learning cycle based on the analytical process can help identify unforeseen areas of public concerns as well as potential barriers for required interventions, thus maximizing vaccine uptake and minimizing health care disparities across demographic communities.[4]

In 2019, WHO named vaccine hesitancy as one of the top 10 threats to global health.[5] The mutable nature of anti-vaccination calls for new modes of analysis to characterize not only the temporal features of hesitancy but also the spatial (e.g., local, regional, national, or international) features and their effects on vaccine uptake. The real-time data on social media also allow investigation into contextual events that can help us understand the barriers to vaccination. This study leverages a multi-level and integrated analytical pipeline, which includes NLP-enabled modeling, time series, and geospatial analyses of social media data. We provide a comprehensive analysis of the attitudes of citizens located in the United States toward the COVID-19 vaccination.

This study aims to analyze the Tweet data after the COVID-19 vaccine deployment from January 2021 to February 2022 and answer three questions: (1) What is the sentiment trend towards COVID-19 vaccination over time; (2) What is the geospatial pattern of the sentiment towards COVID-19 vaccination and over time; (3) What are the popular topics and key barriers towards COVID-19 vaccination?

## METHODS

This study combined machine and human intelligence to perform the analyses. We collected Tweet posts by residents in the United states from the Twitter Application Programming Interface (API) between January 2021 and February 2022. The analyses in this study were mainly conducted based on the Bidirectional Encoder Representations from Transformers (BERT) model, which was used to explore the main sentiment expressed in the Tweets.[6] The longitudinal event analysis allowed us to dive a step further into the sentiment pattern over time and explain some of the fluctuations in the time series results. We also carried out topic modeling focusing on hashtag-based topics. We explored the popular topics from the perspective of sentiment, time series, and geographic pattern, respectively. We also incorporated human intelligence in the analytical process by adding qualitative synthesis of the key barriers and mapping them onto a health care access framework. The overall analytical flow is exhibited in **Figure 1**.

**Figure 1.**
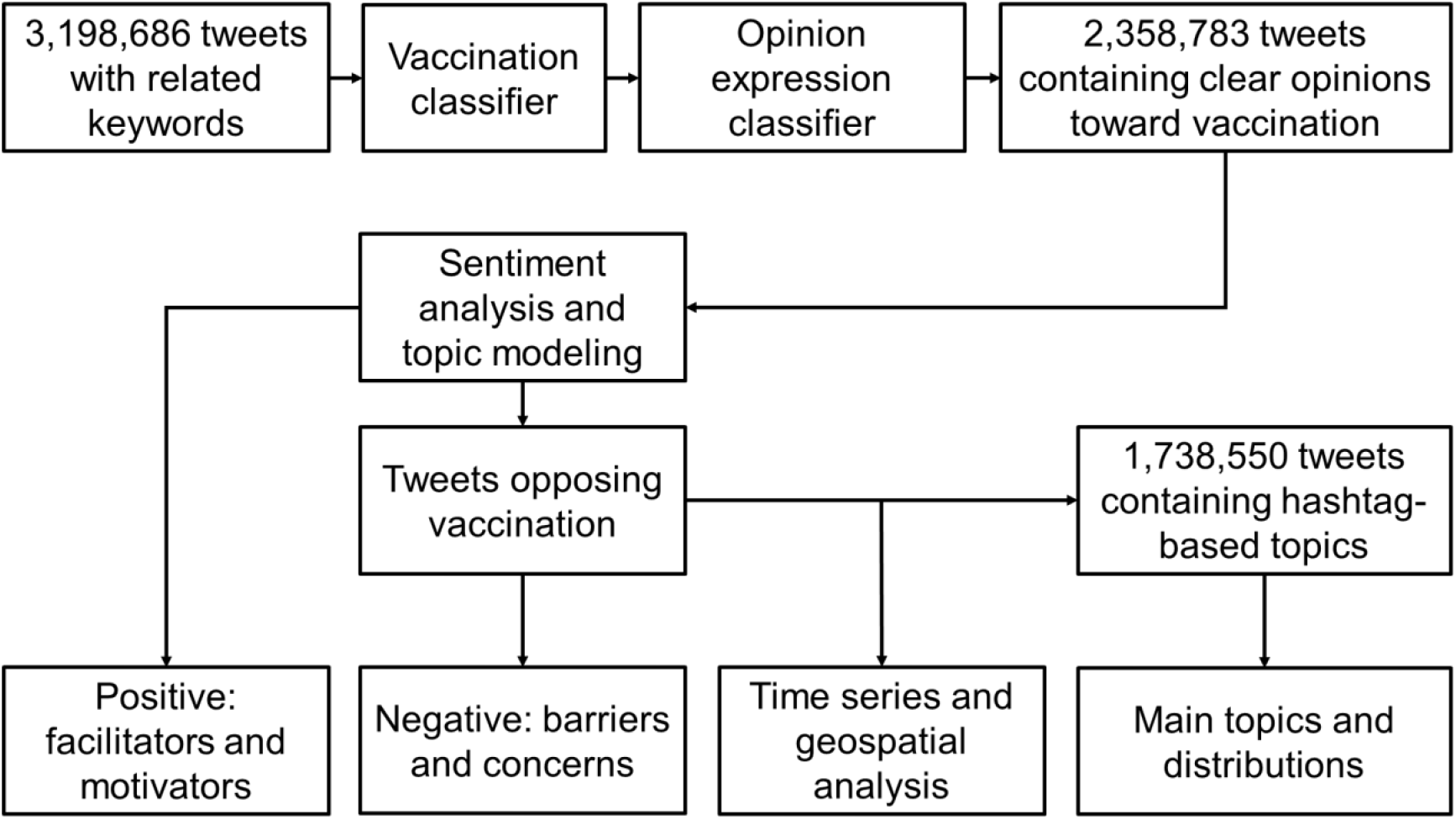
Methods pipeline and flowchart.

Since the data analyzed in this study were completely in the public domain, no ethics review was necessary. We conducted a thorough assessment of the privacy risk that our study posed to individuals to ensure compliance with relevant sections of the General Data Protection Regulation.[7] Further, to comply with privacy laws and social network policies to collect data from Twitter, we did not share or publish personal health information.[8]

### BERT model

BERT is a word representation model that uses unannotated text to perform various NLP tasks such as classification and question answering.[6] By considering the context of a word using the words before or after, we can produce embeddings for words that are more context-aware. This study used the pre-trained sentence BERT model to generate the embedding vectors for emotion classification tasks.

### Sentiment analysis

The sentiment analysis refers to the technique that utilizes NLP and computational linguistics tools to identify or quantify the affective states of the text.[9] Several theories conceptualize emotional states along two dimensions: valence and arousal.[10] Arousal is the level of autonomic activation produced by an event and ranges from calm to excited, while valence describes the level of pleasantness and ranges from negative to positive.[11] For text, arousal states are difficult to recognize, so estimation or classification of valence is the main task of sentiment analysis. Some studies used rule-based approaches such as sentiment dictionaries or traditional supervised classification models such as Logistic Regression, Decision Tree, and Support Vector Machines (SVM) to handle the sentiment analysis task.[12, 13] However, because these methods could not capture contextual information and interdependence among words, their performances were unpromising when it came to long sentences or complicated contexts.[14] Later, Recurrent Neural Network (RNN)-based models were widely applied in text and audio tasks to process sequential data.[15] Recently, with the development of the powerful attention mechanism for dealing with sequential data, transformer-based deep learning models have obtained state-of-the-art results on a range of NLP tasks.[16] Compared to RNNs, transformers are able to capture long-range dependencies better. Even though transformer-based models are harder to be trained from scratch due to a large number of parameters, publicly available pre-trained models, such as BERT and XLNet, enable researchers to easily use transfer learning to train high-performed transformer models on various NLP tasks.[17]

### Hashtag-based topic

Hashtags are central to organizing information on Twitter. Designated by a “hash” symbol (#), a hashtag is a keyword assigned to the information that describes a Tweet and aids in searching. Hashtags organize the discussion around specific topics or events. Hashtag use has become a unique tagging convention to help associate Twitter messages with certain events or contexts.[18] A Twitter hashtag also embodies user participation in the process of hashtag innovation, especially as it pertains to information organization tasks. We leveraged the latent semantic analysis (LSA) [19] and Latent Dirichlet Allocation (LDA) [20] to identify and aggerate the main topics.

### Data Preparation and Pre-processing

We collected the Tweets data from a public Twitter dataset, which contained daily Tweets data related to COVID-19.[21] We used Twitter-API to get raw text Tweets and their corresponding user profiles. We retained Tweets related to COVID-19 vaccines using keywords and removed non-US Tweets based on user location information. Vaccine-related Tweets were collected by detecting keywords on the topic of the vaccine such as “vaccine” and “vaccination”. We also tracked information such as the number of Tweets and followers of each user to identify ghost-writers. After obtaining the vaccine-related Tweets data, we processed the text data by removing irrelevant content for this study such as links. To train and test the sentiment analysis model, we randomly selected 2,500 Tweets for annotation. Then, two classification models based on BERT were trained and selected to filter irrelevant Tweets and predict sentiment states. During the training process, we used back translation for data augmentation.[22] After training, these two classification models would be applied to all the Tweets data.

After data collection, we processed the data by removing redundant contents and potential noises. The specific dataset collection and pre-processing were as following steps:

a. Based on Tweet_id of the public dataset, Tweets data including both text and user profiles were downloaded through Twitter-API.
b. Based on the Tweet language and the user location, the Tweets posted by users in the U.S. were selected.
c. Tweets that did not include vaccine-related words were dropped.
d. To speed up the data collection, massively parallel processing was used to download Tweets and filter irrelevant data.
e. After obtaining the data, the number of daily Tweet posts of each user was calculated locally.

### Data annotation

After obtaining vaccine-related Tweets data, to train a sentiment analysis model, we annotated a total of 2,500 Tweets in the following steps: (1) in order to avoid the bias caused by topics that changed over time, we randomly select 100 Tweets for each month from January 2021 to February 2022 (n=1,400 in total); (2) two authors (J.Y. and J.H.) individually annotated 700 Tweets into four categories: positive, negative, irrelevant, and unsure, (3) after discussion, all the authors checked all annotations and finally annotated these Tweets into three categories: positive, negative, and irrelevant. We only selected the Tweets that expressed a clear sentiment (positive or negative) for the follow-up analyses.

### Data augmentation

Tweet texts sometimes have formats like abbreviations, misspellings, etc. Meanwhile, relying on human intelligence to analyze a large amount of text data and train a robust model would not be feasible. To solve these issues, we used back-translation to augment the annotated data. With the effective application of deep learning translation models in sentence-level translation, translation tools could translate lots of languages into each other with promising performance; and they were also robust to handle typos and abbreviations.[23] We leveraged the Google Translate API to perform this task, which could translate more than 130 languages.[24]

We employed the back-translation strategy in the following steps. First, we categorized all the available languages into language families based on information provided by Wikipedia. According to the populations of native speakers of each language family, we further selected five intermediate languages (Chinese, German, French, Russian, and Japanese) of the most used language families. Finally, a Tweet was translated into the target intermediate languages iteratively for five rounds, and the translated sentences were then translated back into English. To improve training efficiency, the back-translation was done before training and all data was saved locally. By randomly selecting 500 samples for manual verification, we found that back-translation not only enriched vocabulary and syntax, but also addressed the problems of colloquial expressions and typos. During the training process, we switched original Tweets and back-translated Tweets with different intermediate languages for each epoch. According to the test results, the back-translation improved the model performance (F1-score) by more than 5%.

### Model Training and Applications

As the labeled dataset by human intelligence was not sufficient to support the training of the sentiment from the scratch, we took advantage of transfer learning to complete the task. In the earlier NLP research, pre-trained language models were usually used as feature extractors to obtain vector representations of words; then machine learning models were trained with these embeddings. Later, with the remarkable breakthroughs in deep learning models, the parameters of a pre-trained model could be fine-tuned by re-training the model on a downstream task. Recently, transformer-based models like BERT achieved state-of-the-art performances on different kinds of down streaming tasks such as text classification and sentiment analysis.[6] To transfer the pre-trained model to the sentiment task, we modified the torch-version pre-trained BERT model provided by Hugging Face by using the [CLS] token as input to a fully connected network with one hidden layer, and the softmax activation function as the last layer of the model to perform classification.[25] The analysis task was divided into two binary classification tasks: irrelevant content detection and sentiment analysis (**Figure 2**). The [CLS] was used to capture global features of the whole input text, and the [SEP] token helped the model to separate sentences. The E. denotes the embedding of each word, and the B. denotes the feature encoded by the BERT encoder.

**Figure 2.**
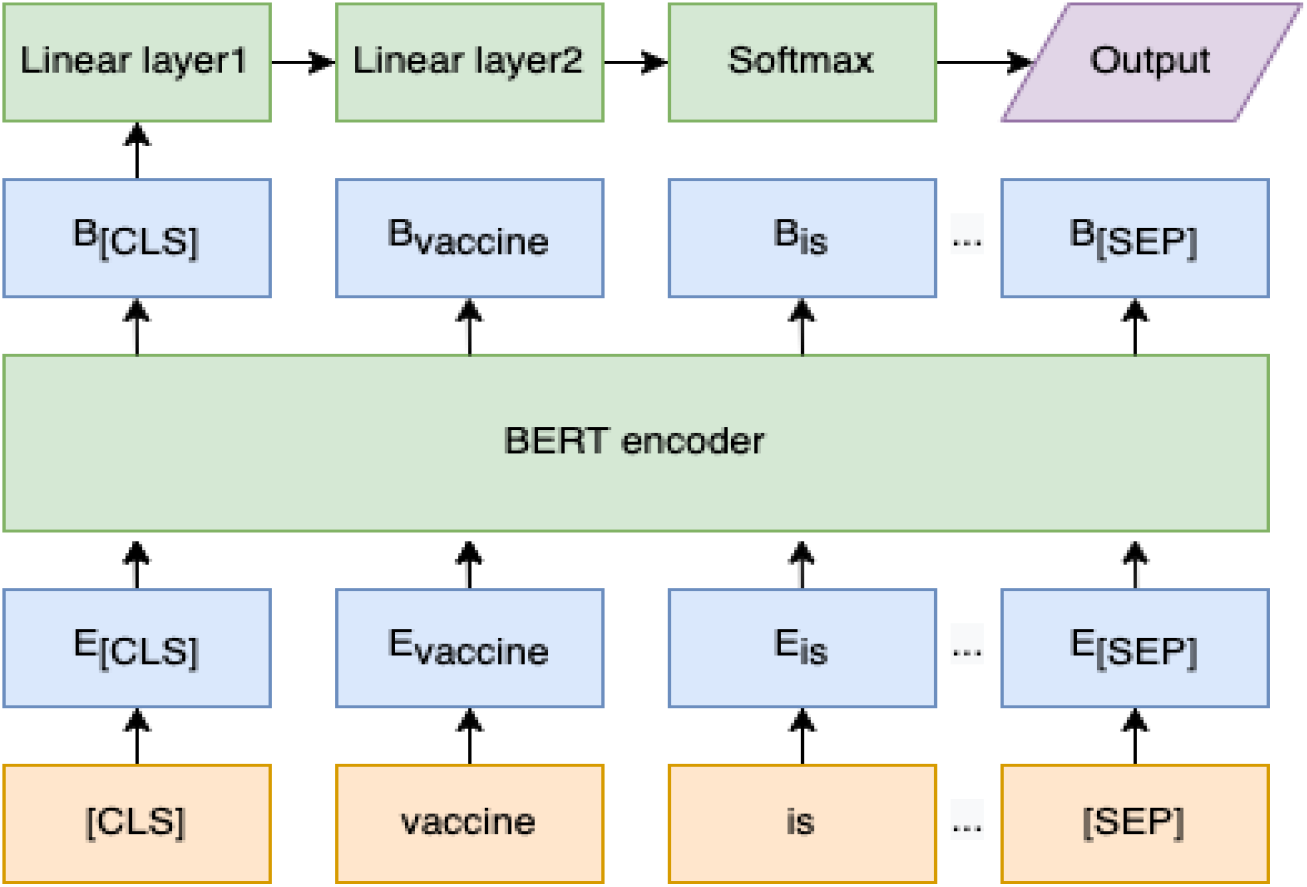
The framework of the sentiment analysis model.

We conducted our experiments on an NVIDIA GTX1070 TI GPU. The hyperparameters settings for the training were:

a. Epochs = the number of back-translation languages * 3
b. Batch size = 4
c. Learning rate = 1e^-5^
d. Weight decay = 1e^-5^
e. Max sequence length = 512

As our annotated data might not be balanced, we considered 80% of each category as training data to fine-tune the weights of the pre-trained model, 10% as validation data to select the best-performed model for the training, and 10% as testing data to evaluate the performance of the final models. Finally, the irrelevant content detection achieved 81.26% accuracy, and the sentiment analysis model achieved 79.67% accuracy and 87.53% F1 score on the testing dataset.

Next, we applied these two training models to the whole dataset. We filtered Tweets that were predicted as irrelevant content. The rate of Tweets with a negative sentiment each day was then calculated. We utilized the location information of users to evaluate the geospatial variations in public sentiment among the states and temporal variations in public sentiments toward COVID-19 vaccination. We identified, evaluated, and associated the key events that impacted the positive or negative sentiments to the temporal trends. We also conducted qualitative syntheses of Tweets on points of interest to identify underlying themes and validated insights from the Tweets.

## RESULTS

A total of 3,198,686 Tweets met our inclusion criteria. After applying the vaccination classifier model and opinion expression classifier model, 839,903 Tweets that were not relevant to COVID-19 vaccination or expressed an unclear sentiment were removed. The remaining 2,358,783 Tweets were then analyzed using sentiment analysis and topic models. Among the 2,358,783 Tweets, 824,755 (35.0%) expressed negative opinions towards vaccination while 1,534,028 (65.0%) demonstrated positive opinions. We further identified 1,738,550 (73.7%) Tweets that had hashtags (i.e., “#”) within their content, which was used to analyze the topic.

**Figure 3** demonstrates the weekly rolling average time-series results of Twitter sentiments (red line) and vaccinations (green line). Because some states did not administer vaccines during the weekend, we used the weekly rolling average rather than daily measures. Overall, the two lines present opposite tendencies over time. There was a steady increase in vaccination since January 2021 and reached the peak in April 2021 and followed by a sharp decrease after April 2021. There are some potential reasons for this pattern: (1) In April 2021, the U.S. surpassed 200 million vaccinations administered, most of whom might hold positive sentiment toward vaccination; individuals who posted Tweets related to vaccines might be the remaining population who held negative sentiment; (2) CDC and FDA paused the use of the Johnson and Johnson COVID-19 vaccine because of the blood clot complications;[26] this event might increase individuals’ concerns about vaccine safety; (3) In June 2021, the Delta variant, which was first identified in India in late 2020, became the dominant variant in the U.S. The variant kicked off a third wave of infections during the summer of 2021, which might cause the dramatically decreased vaccination. Since the fall of 2021, many states issued vaccine mandates policy. This policy likely increased the vaccinations between October 2021 and December 2021. In the middle of December 2021, the number of daily vaccinations deceased, and the reasons might be: (1) the number of fully vaccinated has reached a saturated state; according to the real-time data, more than 83% of individuals had at least 1 dose and more than 71% individuals were fully vaccinated.[27] (2)The first U.S. case of COVID-19 attributed to the Omicron variant was detected in December 2021,[28] which might cause the increase pattern like the Delta variant.

**Figure 3.**
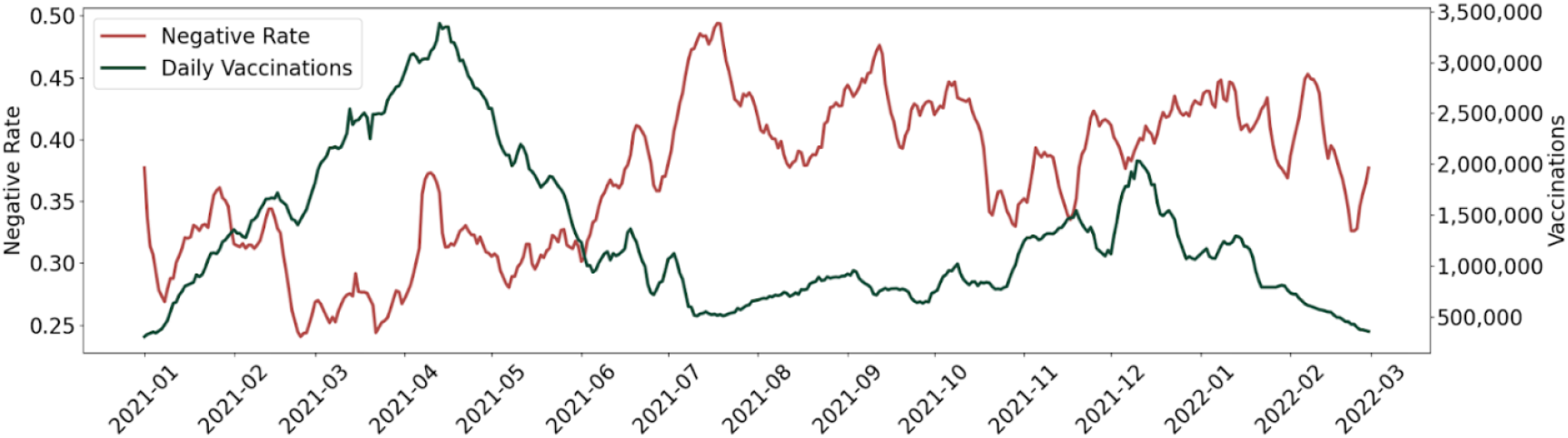
Weekly rolling average time-series results of Tweets sentiments (left Y-axis) and vaccinations (right Y-axis).

**Figure 4** presents the geospatial mapping of social media public negative sentiments in the United States toward the COVID-19 vaccine. A geospatial map of overall (averaged) sentiments at the state level indicates that most states had a moderate negative sentiment. The states with relatively higher negative sentiment toward COVID-19 vaccination were concentrated in the west, and some states in the east and southeast regions, the top five states of negative sentiment include Wyoming, Pennsylvania, Florida, Hawaii, and California.

**Figure 4.**
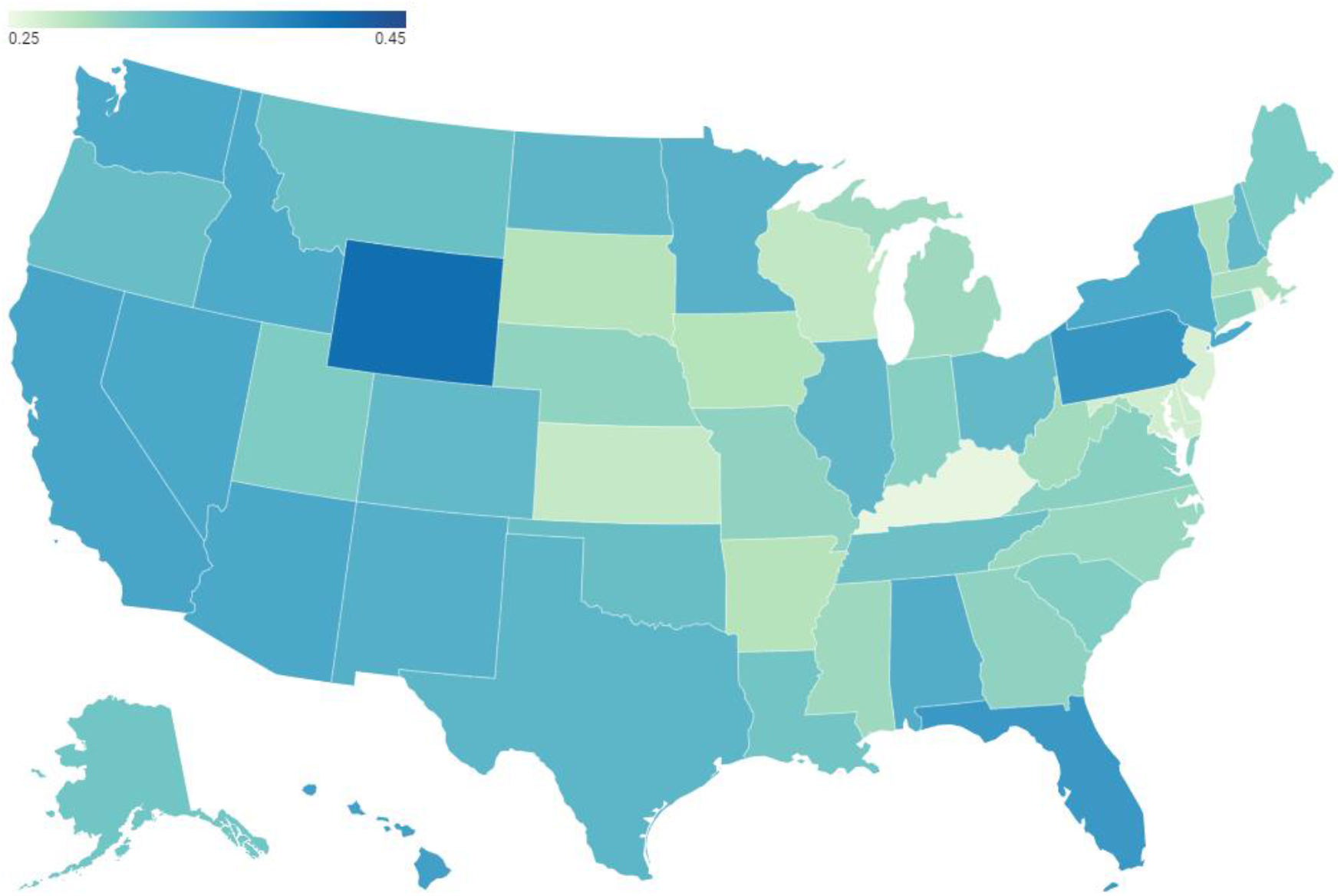
Geospatial mapping of social media public negative sentiments in the United States toward the COVID-19 vaccination.

**Figure 5** demonstrates the public negative sentiments in each state of the United States toward the COVID-19 vaccine over time. As time went by, the negative sentiment rate increased and reached the highest in July 2021, which aligned with the results in **Figure 2**. We also see variabilities across the states.

**Figure 5.**
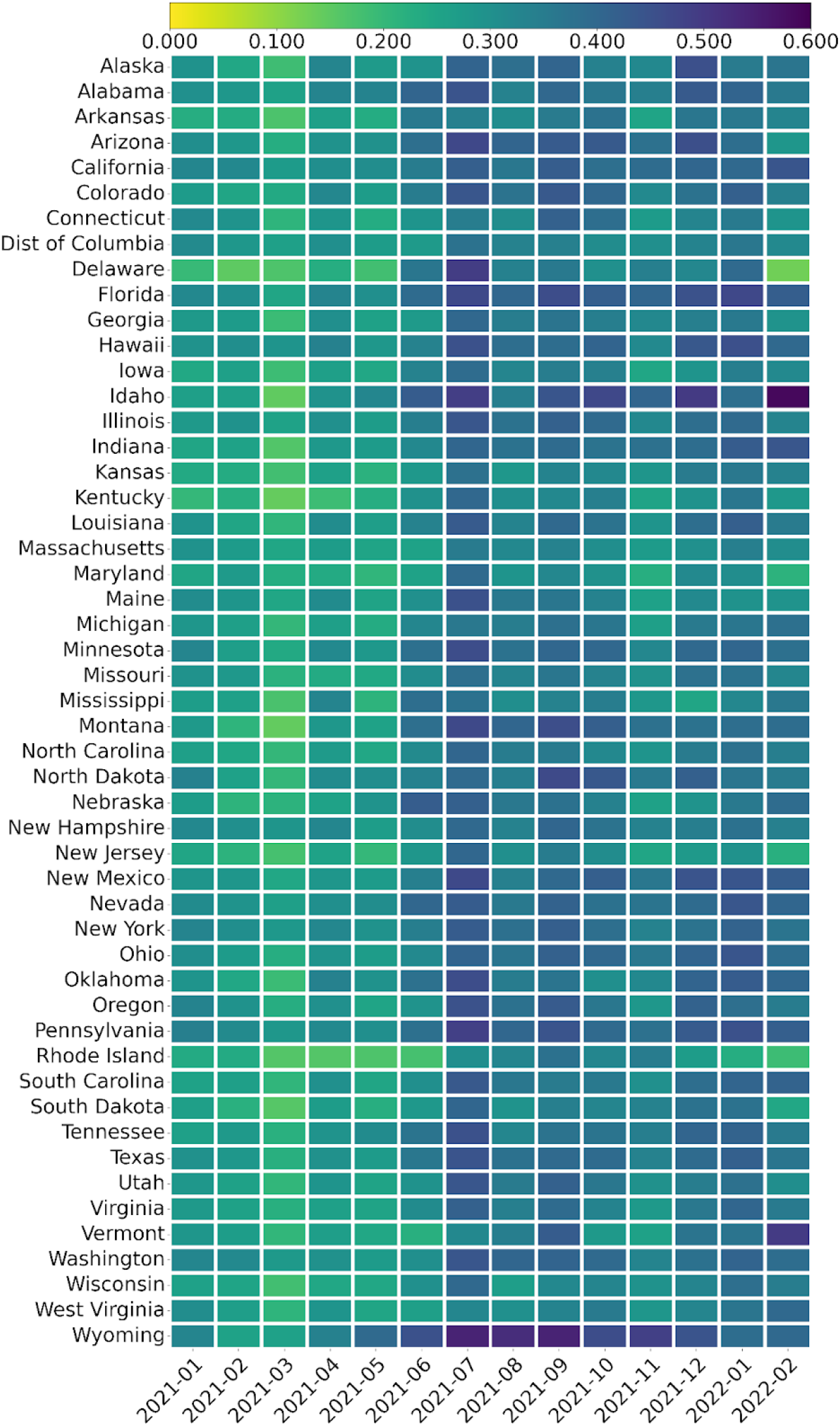
Social media public negative sentiments in each state of the United States toward the COVID-19 vaccination over time (sorted by alphabetical).

**Figure 6** demonstrates major word rates in each state of the United States regarding the COVID-19 vaccine. The numerator is the word’s frequency in the state and the denominator is the total number of Tweets in the state. The top ten words are Pfizer, first, fully, today, cases, unvaccinated, shot, Johnson, available, and children. We see moderate variation across the states, but some words were substantial in some states. For example, “available” has a high rate in Florida; “first” and “available” have high rates in Kentucky; “fully”, “today”, and “cases” have high rates in Maine, Rhode Island, and Utah.

**Figure 6.**
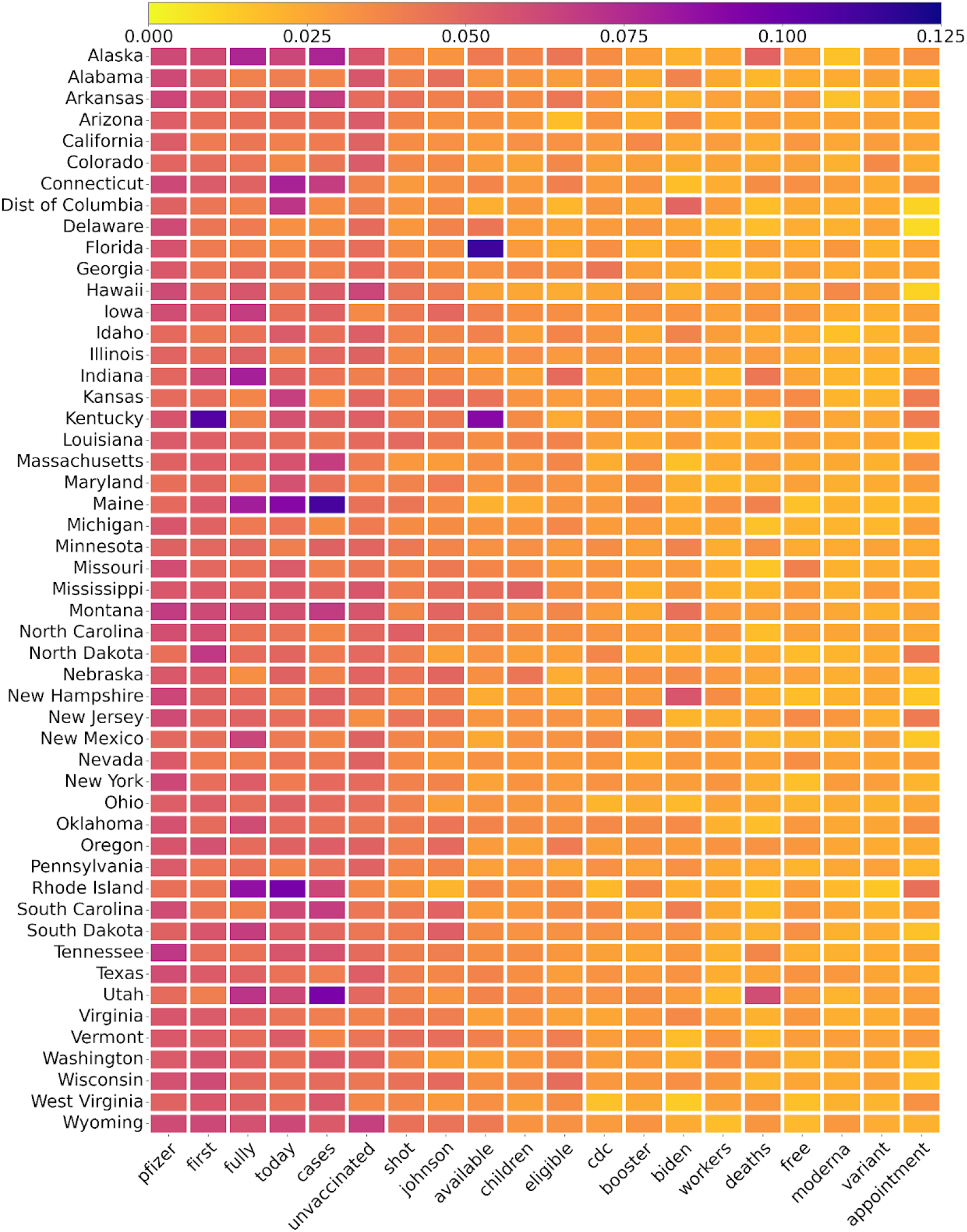
Major words rate on the social media in each state of the United States toward the COVID-19 vaccination (sorted by alphabetical). Numerator: the word’s frequency in the state; denominator: the total number of Tweets in the state.

**Figure 7** demonstrates the major hashtag-based topic rates on social media in each state of the United States toward COVID-19 vaccination. The numerator is the topic’s frequency in the state and the denominator is the total number of Tweets in the state. Among the selected 18 topics, Wisconsin has the highest total rate, which means the COVID-19 vaccine-related Tweets that residents in Wisconsin posted contained most of these selected topics. In addition, the majority of relevant Tweets in Wisconsin contained “thisisourshot”, which meant this campaign gained a good buy-in in Wisconsin. The top 10 topics are Pfizer, breaking, wearamask, smartnews, moderna, publichealth, cdc, omicron, thisisourshot, and wecandothis. Only “Pfizer” is the only same topic as the results of the major words rate in **Figure 6**, which means the Pfizer vaccine gained the most popularity on the Twitter platform.

**Figure 7.**
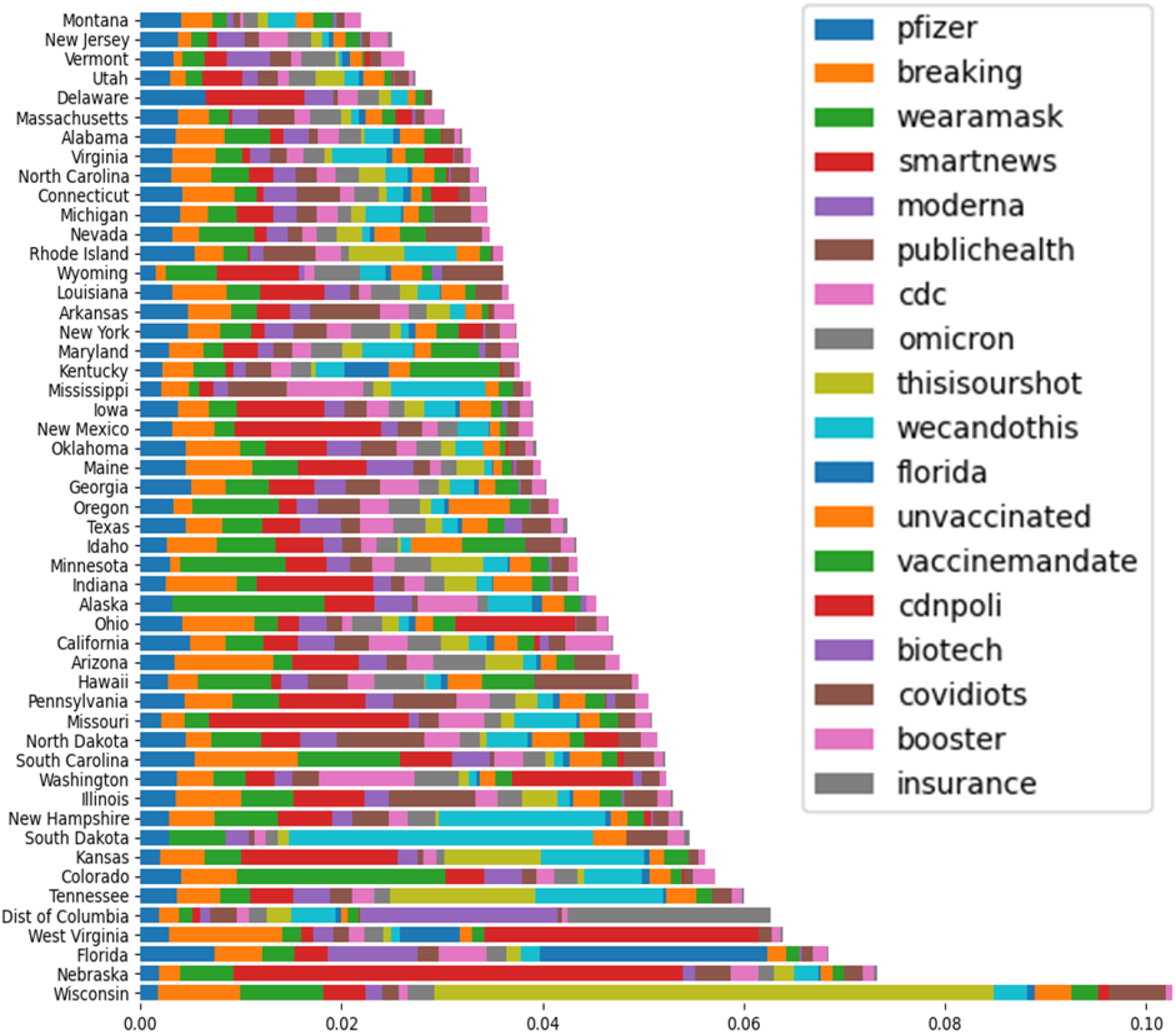
Major hashtag-based topic rate on the social media in each state of the United States toward the COVID-19 vaccination (sorted by topics’ rate).

**Table 1** demonstrates major categories of reasons or concerns for opposing COVID-19 vaccination. We classified the 2,500 randomly selected Tweets, discussed the themes, and mapped them on Levesque’s model,[29] which was designed to explain the comprehensiveness and dynamic nature of access to health care with five domains of accessibility (Approachability, Acceptability, Availability and accommodation, Affordability, and Appropriateness). We further illustrated the themes and provided examples within each of the domains. The main barrier themes include accessibility, hesitancy, dislike forcing, safety concern, mistrust, manufacturing delays, inequity, conspiracy theory, and misinformation. Among the barriers, mistrust (31.8%), hesitancy (27.9%), and safety concern (20.3%) are the top three themes. Some individuals declined in trust of expertise and authority, and different modes of belief-based extremism.[30] Political polarization, as well as libertarian views and alternative health care advocacy, triggered public questioning about the importance, safety, and effectiveness of COVID-19 vaccines.[31] In addition, manufacturing delays also increased the negative sentiment toward vaccination.[32] For example, in February 2021, vaccine distribution was disrupted in several states, including Texas, Missouri, Alabama, and New Hampshire due to severe winter storms.[33]

**Table 1.**
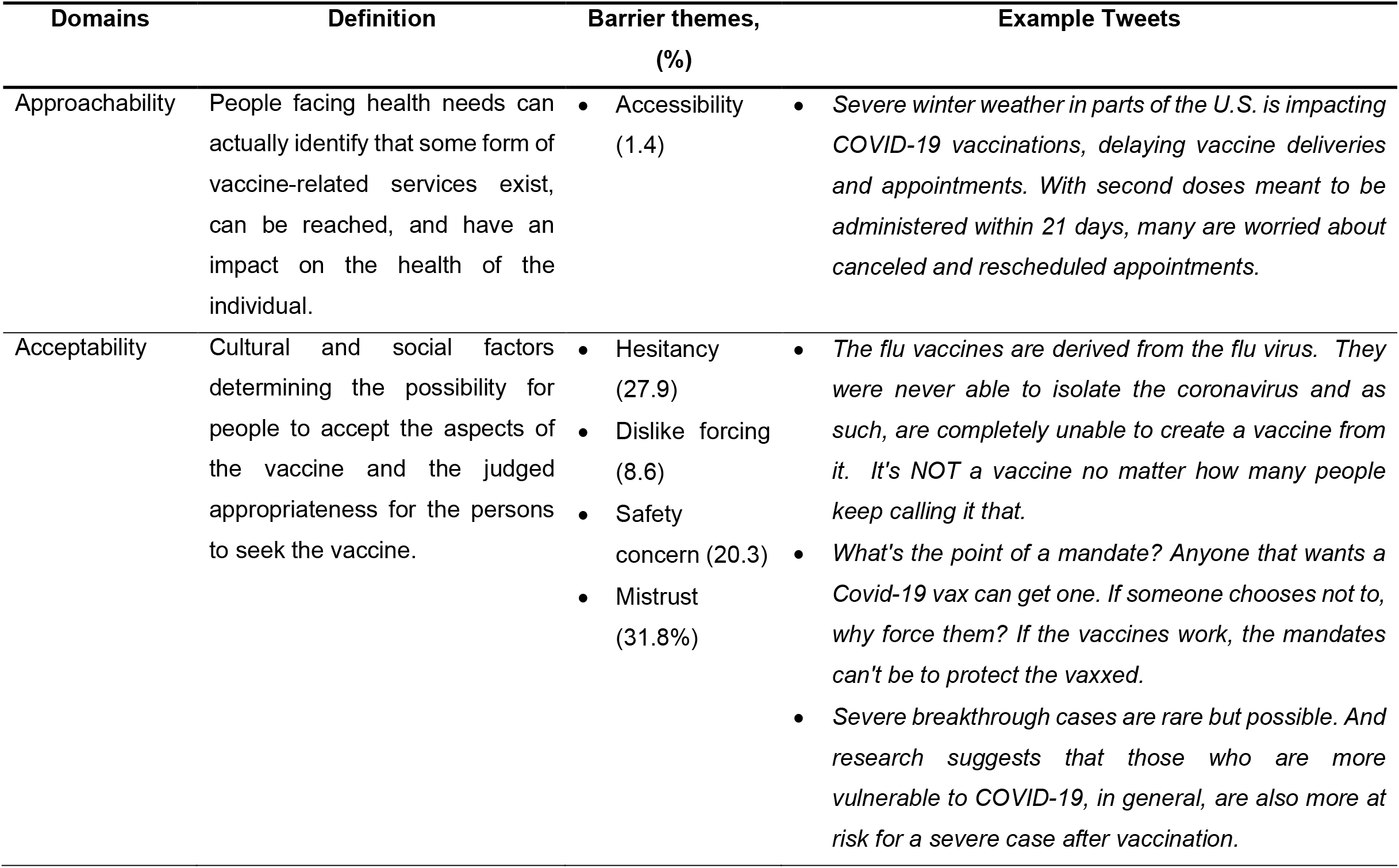
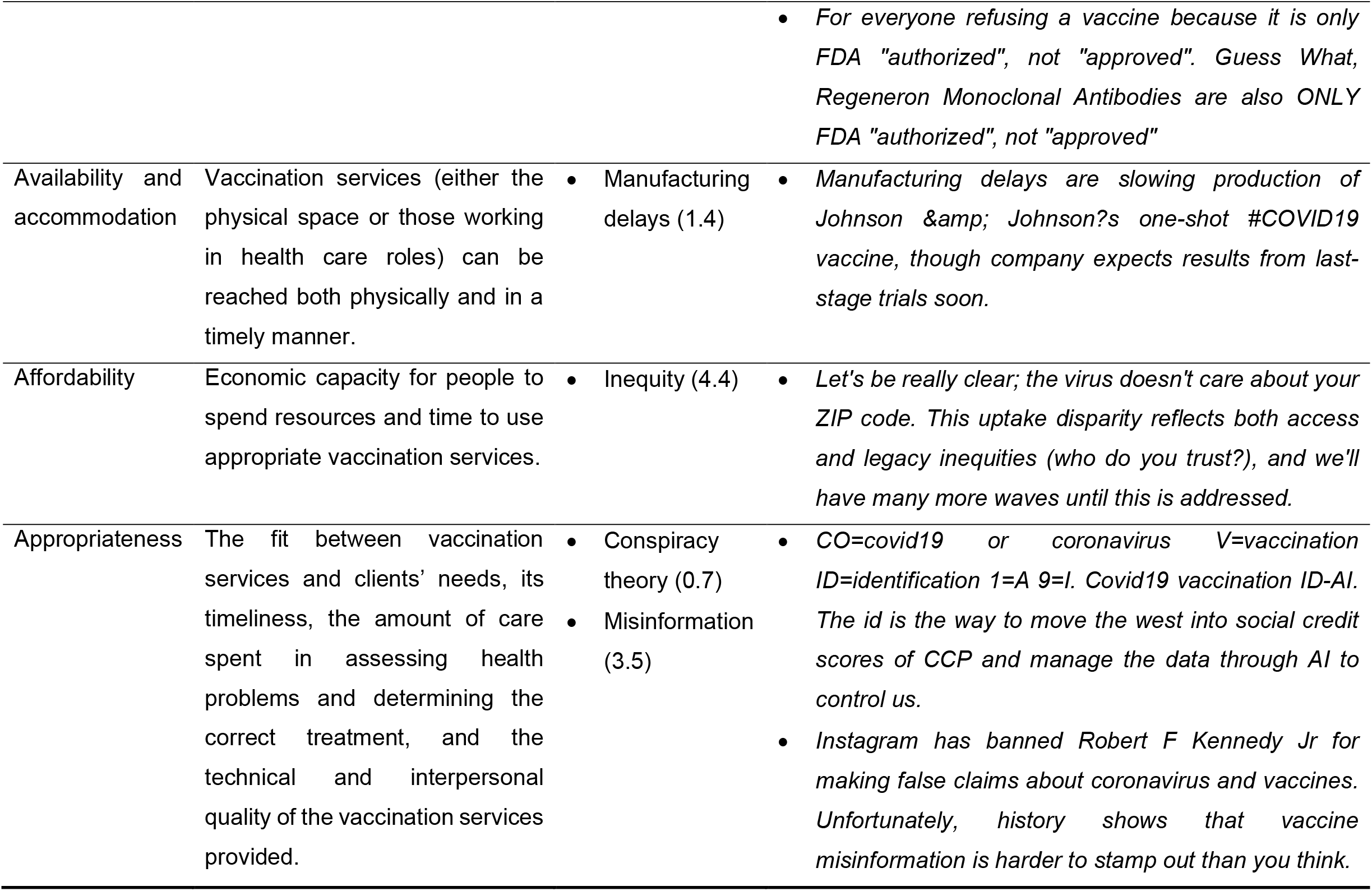
A pragmatic taxonomy of reasons or concerns for opposing COVID-19 vaccine or vaccination hesitancy based on the Levesque’s model.

| Domains | Definition | Barrier themes,<br>(%) | Example Tweets |
| --- | --- | --- | --- |
| Approachability | People facing health needs can actually identify that some form of vaccine-related services exist, can be reached, and have an impact on the health of the individual. | • Accessibility (1.4) | • *Severe winter weather in parts of the U.S. is impacting COVID-19 vaccinations, delaying vaccine deliveries and appointments. With second doses meant to be administered within 21 days, many are worried about canceled and rescheduled appointments.* |
| Acceptability | Cultural and social factors determining the possibility for people to accept the aspects of the vaccine and the judged appropriateness for the persons to seek the vaccine. | • Hesitancy (27.9)<br>• Dislike forcing (8.6)<br>• Safety concern (20.3)<br>• Mistrust (31.8%) | • *The flu vaccines are derived from the flu virus. They were never able to isolate the coronavirus and as such, are completely unable to create a vaccine from it. It's NOT a vaccine no matter how many people keep calling it that.*<br>• *What's the point of a mandate? Anyone that wants a Covid-19 vax can get one. If someone chooses not to, why force them? If the vaccines work, the mandates can't be to protect the vaxxed.*<br>• *Severe breakthrough cases are rare but possible. And research suggests that those who are more vulnerable to COVID-19, in general, are also more at risk for a severe case after vaccination.* |
|  |  |  | • *For everyone refusing a vaccine because it is only FDA "authorized", not "approved". Guess What, Regeneron Monoclonal Antibodies are also ONLY FDA "authorized", not "approved"* |
| Availability and accommodation | Vaccination services (either the physical space or those working in health care roles) can be reached both physically and in a timely manner. | • Manufacturing delays (1.4) | • *Manufacturing delays are slowing production of Johnson & Johnson's one-shot #COVID19 vaccine, though company expects results from last-stage trials soon.* |
| Affordability | Economic capacity for people to spend resources and time to use appropriate vaccination services. | • Inequity (4.4) | • *Let's be really clear; the virus doesn't care about your ZIP code. This uptake disparity reflects both access and legacy inequities (who do you trust?), and we'll have many more waves until this is addressed.* |
| Appropriateness | The fit between vaccination services and clients' needs, its timeliness, the amount of care spent in assessing health problems and determining the correct treatment, and the technical and interpersonal quality of the vaccination services provided. | • Conspiracy theory (0.7)<br>• Misinformation (3.5) | • *CO=covid19 or coronavirus V=vaccination ID=identification 1=A 9=I. Covid19 vaccination ID-AI. The id is the way to move the west into social credit scores of CCP and manage the data through AI to control us.*<br>• *Instagram has banned Robert F Kennedy Jr for making false claims about coronavirus and vaccines. Unfortunately, history shows that vaccine misinformation is harder to stamp out than you think.* |

## DISCUSSION

Social media like Twitter provides the opportunity to collect data related to vaccination in nearly real-time. This digital platform also allows new methods of analysis and the opportunity to investigate the effect of the sentiment on vaccine uptake.[34] In this study, we analyzed public opinions expressed on Twitter regarding COVID-19 vaccination in the United States. We studied a total of 3,198,686 Tweets collected from January 2021 to February 2021 using an analytical strategy that combined qualitative content analysis [35] for understanding opinions expressed in subtle human language and machine learning for scalability. Comparative analysis revealed that over the 14-month study period, the overall public sentiment toward COVID-19 vaccination was moderate positive. The sentiment across the states showed manifest variability. The top three barriers to vaccination were mistrust, hesitancy, and safety concerns.

The early detection of an opinion shift might be useful in the context, in which many countries from around the world are working on the COVID-19 vaccination, as it would promote actions aimed at increasing the general public’s confidence towards vaccination. Many people search online for health-related information and the information will impact patient decision-making. It is therefore essential to understand what is shared online.[36] The results demonstrate the potential of NLP-based real-time social media monitoring of public sentiments and attitudes to help detect and prevent vaccination hesitancy and concerns. This monitoring may inform more effective strategies for vaccine deployment, under conditions of uncertainty, including decisions on prioritization and equitability, to help maximize the uptake of the vaccines.[35]

Given the dramatic changes in the communication landscape that fuel the rapid spread of vaccine information alongside misinformation, new methodologies are needed to monitor emerging vaccine concerns over time and place in order to better inform appropriate responses.[37, 38] We analyzed the temporal variations in public sentiments toward COVID-19 vaccines in the United States. We identified, evaluated, and mapped the keywords and hashtag-based topics impacting positive and negative sentiments to the temporal trends. Mapping vaccine hesitancy at a local level is also one important step toward addressing it, along with other needed interventions at the individual and community levels. We mapped spatial variations in public sentiment to regions in the United States. The geospatial maps could help identify areas with more negative sentiments toward COVID-19 vaccination, which can be further studied for potential interventions to allay the underlying public concerns.[39] We found that different states presented various trend patterns in sentiment change over time. Geospatial analysis and map visualization better portray more aspects of residents’ attitudes towards COVID-19 vaccination, which may be helpful for government and public health agencies to conduct COVID-19 vaccination campaigns in areas that need more attention and efforts to address the barriers and concerns. The large volume of timely data on social media has provided an opportunity to develop spatially detailed estimates of vaccination sentiment (i.e., mapping by location). Spatially refined estimates of vaccination sentiment have proved to be useful in local efforts to increase vaccination rates.[40] The information may be used by community-based programs to tailor their efforts to local areas that have the greatest need.[41] The geographical patterns can also be used to identify places to provide mobile vaccination clinics and initiate measures for reducing barriers to vaccination. Local information can also be used to monitor the effectiveness of local interventions, including the effect of various types of vaccination mandates. Furthermore, there have been cases where vaccine debates have been purposefully polarized, thus exploiting the doubting public and system weaknesses for political purposes, while waning vaccine confidence elsewhere may be influenced by a general distrust in the government and scientific elites.

Future work should develop diverse and effective machine learning classifiers to facilitate opinion mining using social media data and the automatic and continuous extraction and monitoring of public opinions. Large and complex datasets on vaccination should also be analyzed according to other identifiers such as ZIP Code and individual characteristics, including social determinants of health, which can help to advance further micro-target vaccine deployment efforts.[42] This information is also potentially critical for monitoring progress toward vaccination equity. In addition, future work should include more diverse social media platforms representing different types of user groups, different interaction modalities, and geographic settings to address health care disparity.

## CONCLUSION

This study demonstrates the potential of an analytical pipeline, which integrates NLP-enabled modeling, time series, and geospatial analyses of social media data. Through the analysis of a large Twitter dataset using a combination of natural language processing and qualitative content analysis, we classified the public’s attitude toward COVID-19 vaccination, the temporal trend over time, and geographic sentiment distribution. The results showed that while generally more Tweets held positive opinions on vaccination, negative opinions were not uncommon. The sentiment towards vaccination across the states showed manifest variability. The top three barriers to vaccination were mistrust, hesitancy, and safety concerns. The resilience of vaccination programs may be influenced by the rapid and global spread of misinformation. Public confidence in COVID-19 vaccines can be exacerbated by unproven concerns regarding vaccine safety, which seed doubt and mistrust. The NLP-enabled real-time social media monitoring of public sentiments and attitudes can help detect public sentiment towards COVID-19 vaccination, which may help solution providers to understand the reasons why some social groups may be reluctant to be vaccinated against COVID-19. The results could provide support for developing tailored policies and communication strategies to facilitate COVID-19 vaccination.

## Data Availability

All data produced in the present work are contained in the manuscript.

## AUTHOR CONTRIBUTIONS

J.Y. conceived and designed the study, contributed to the analyses, and drafted the manuscript. J.H. contributed to the analyses. All the authors contributed to the interpretation of the results and revision of the manuscript. All the authors read and approved the final version of the manuscript.

## DATA AVAILABILITY STATEMENT

All data referred to in the manuscript are publicly available.

